# Enhanced comparisons of COVID-19 mortality across populations

**DOI:** 10.1101/2020.06.14.20131318

**Authors:** Chalapati Rao, Suhail A. Doi, Gail Williams

**Affiliations:** Research School of Population Health, Australian National University, 62, Mills Road, Action ACT 2601, Australia, Email; Department of Population Medicine, College of Medicine, Qatar University, Doha, Qatar; School of Public Health, University of Queensland, 288, Herston Road, Herston QLD 4006, Australia

## Abstract

**Background:** The reported crude case fatality rate (CFR) for COVID-19 varies considerably across countries. Crude CFRs could by biased by larger proportions of older COVID-19 cases in population data, who are also at increased mortality risk. Such distorted age case structures are a common feature of selective COVID 19 testing strategies in many countries, and they potentially mask underlying differences arising from other important factors such as health system burden.

**Methods:** We used the method of direct case-age standardisation to evaluate the effects of age variations on CFRs. Data on cases and death by age from Italy, Spain, China, Australia and South Korea were analysed to derive standardised CFRs. Findings were compared across different case age distribution references as standards.

**Results:** Using the South Korean case age distribution as a standard, the fivefold higher crude CFR for Italy is reduced to less than two-fold after adjustment, while the crude CFR difference for Spain is virtually eliminated. The adjusted CFR for Australia is the lowest among all countries.

**Discussion:** Mortality differences based on crude CFRs are exaggerated by age structures, which are effectively controlled by case age standardization. Residual CFR differences could be attributed to health and health system factors. The South Korean case age distribution is an appropriate reference standard, given its robust case detection and contact tracing program. Till reliable population level indicators of incidence and mortality are available, the age-standardized CFR could be a viable option for international comparison of the impact of the COVID 19 epidemic.

**Summary:** *The known:* There are intense debates around the magnitude of and reasons for wide variations in observed case fatality rates (CFRs) from COVID 19 across countries. Age is commonly speculated as a reason, but this has not been technically quantified or explained.

*The new:* The technique of direct standardization using reference distributions of case age structures eliminates the effects of age on CFR, thus enhancing the comparability as well as understanding of differentials

*The implications:* Residual differences between adjusted CFRs can be used to infer health and health system factors that influence mortality in COVID 19 cases in different populations

## Introduction

As the COVID-19 pandemic matures across the world, the case-fatality rate (CFR) has been the most readily measureable mortality indicator from available data. Widely differing reports of CFRs have triggered intense debate in regard to the relative impact of the disease at national level. An international comparison across 25 countries which had reported at least 1000 cases as at 23 March 2020 showed CFRs ranging from 0.38% in Germany to 9.26% in Italy.(1) There is a common understanding among researchers of the inherent bias in the reported numbers of cases and deaths from all populations. (2, 3) Numerators could be biased from different case definitions, varied COVID 19 testing strategies that result in undetected cases, and variations in validity of different COVID tests. Definitions of COVID 19 deaths could vary according to availability of laboratory evidence, or from differences in clinical opinion on the causal or associative relationship between COVID infections and death. (4) Changes in the pace and momentum of the epidemic can also affect CFRs. An attempt has been made to estimate CFR with the denominator based on only closed cases within a cohort, but this was countered by an alternate analysis which demonstrated that the CFR is best measured using total cases as the denominator.(5, 6) During infectious disease outbreaks, CFRs could be affected by biases in both directions, making it difficult to reliably interpret this indicator for comparison across populations. (7)

Increased COVID 19 mortality at older ages has been observed in all populations. Higher CFRs in Italy and Spain have been attributed to their higher proportions of the elderly. (1) However, CFRs are computed from cases rather than the general population itself, and there is insufficient reason to believe that there is increased general risk of exposure to infection among the elderly. Hence, higher crude CFRs in populations such as Italy are likely due to higher rates of case detection among the elderly who are hospitalised for acute or terminal care of COVID 19 disease. (8) Therefore, crude CFRs are not directly comparable across populations, due to different case age structures. In this article, we present an analysis of CFRs adjusted for age distributions of cases, and propose that the age-adjusted CFRs should be used for comparing the impact of the pandemic across countries. We also discuss the justification for this approach, and the broader implications of CFRs as an epidemiological indicator and also as a measure to guide clinical care modalities to improve case outcomes.

## Data and methods

Our analysis is based on the principle of age-standardization, which is a technique to eliminate the effect of age-structure of at-risk populations on the overall rate of outcome of interest. Data on age-distributions of COVID 19 cases and deaths were sourced for Italy, Spain, China, Australia and South Korea (9-14). All data on cases and deaths were available for both sexes together, and for identical age groups.

For each study population, age-specific case fatality rates were computed as proportions of reported deaths out of cases in each age group, along with the crude case fatality rate across all ages. We followed the method of direct standardization, in which the proportional case distribution by age from each country was serially used as a weight to derive the weighted average case fatality rate for each of the study populations, which equals the standardised case fatality rate. The variance for each standardised fatality rate was used to derive a 95 % confidence interval to assess uncertainty around the standardised estimate.

The standardised fatality rate and its variance are given by (15):

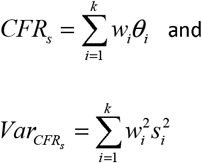

Where *i* = 1,…, *k* indexes age-groups, *w* is the case-standard age-category proportional weight (that sums to 1) and *θ* and *s*^2^ are the age specific CFR and its variance respectively.

## Results

Table 1 shows the age distributions of cases and deaths for all study populations. The data for China include reported cases up to 11 February 2020; while the data for the other countries are up till 30 April, 2020. As expected, age-specific CFRs increase with age in all populations. There is considerable variation in crude CFRs, with the crude rates in Spain and Italy estimated to be approximately three and five times the rate in South Korea.

**Table 1:**
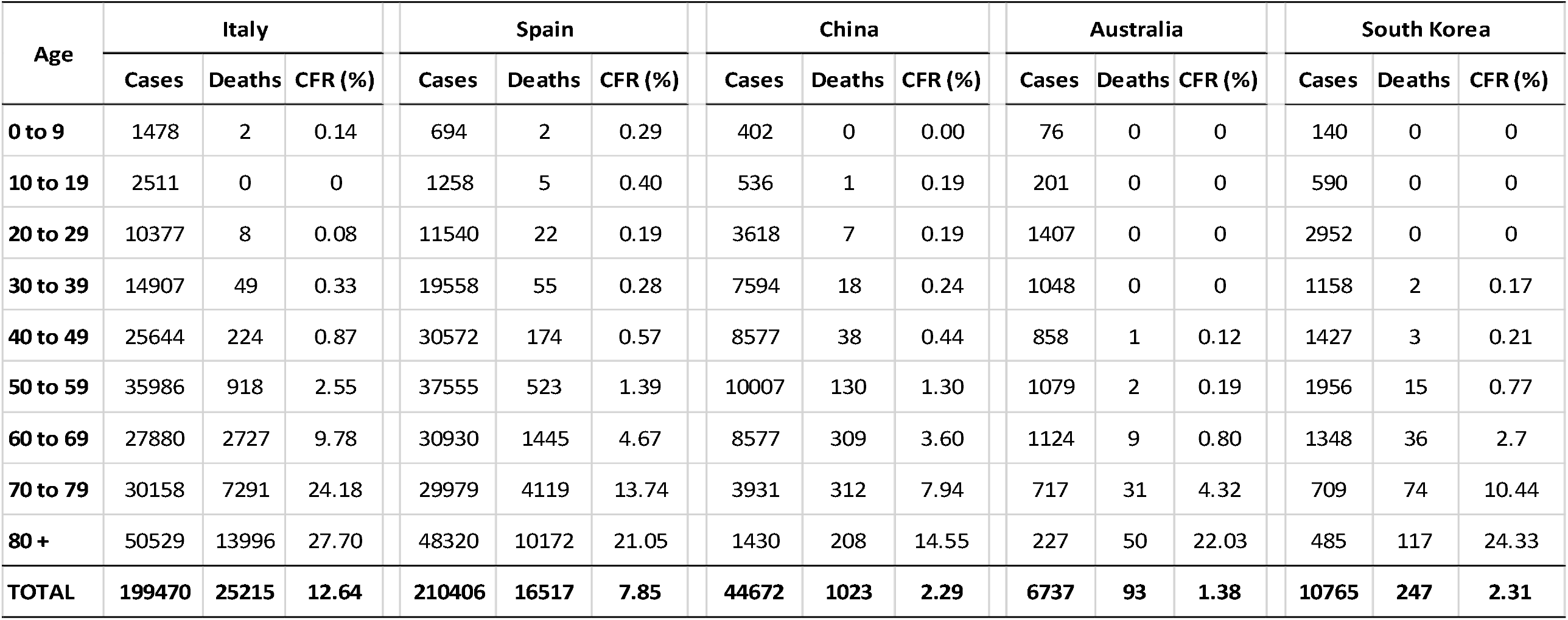
Age distributions of reported COVID 19 cases, deaths, and age-specific case fatality rates from different countries, 2020

| Age | Italy |  |  | Spain |  |  | China |  |  | Australia |  |  | South Korea |  |  |
| --- | --- | --- | --- | --- | --- | --- | --- | --- | --- | --- | --- | --- | --- | --- | --- |
|  | Cases | Deaths | CFR (%) | Cases | Deaths | CFR (%) | Cases | Deaths | CFR (%) | Cases | Deaths | CFR (%) | Cases | Deaths | CFR (%) |
| **0 to 9** | 1478 | 2 | 0.14 | 694 | 2 | 0.29 | 402 | 0 | 0.00 | 76 | 0 | 0 | 140 | 0 | 0 |
| **10 to 19** | 2511 | 0 | 0 | 1258 | 5 | 0.40 | 536 | 1 | 0.19 | 201 | 0 | 0 | 590 | 0 | 0 |
| **20 to 29** | 10377 | 8 | 0.08 | 11540 | 22 | 0.19 | 3618 | 7 | 0.19 | 1407 | 0 | 0 | 2952 | 0 | 0 |
| **30 to 39** | 14907 | 49 | 0.33 | 19558 | 55 | 0.28 | 7594 | 18 | 0.24 | 1048 | 0 | 0 | 1158 | 2 | 0.17 |
| **40 to 49** | 25644 | 224 | 0.87 | 30572 | 174 | 0.57 | 8577 | 38 | 0.44 | 858 | 1 | 0.12 | 1427 | 3 | 0.21 |
| **50 to 59** | 35986 | 918 | 2.55 | 37555 | 523 | 1.39 | 10007 | 130 | 1.30 | 1079 | 2 | 0.19 | 1956 | 15 | 0.77 |
| **60 to 69** | 27880 | 2727 | 9.78 | 30930 | 1445 | 4.67 | 8577 | 309 | 3.60 | 1124 | 9 | 0.80 | 1348 | 36 | 2.7 |
| **70 to 79** | 30158 | 7291 | 24.18 | 29979 | 4119 | 13.74 | 3931 | 312 | 7.94 | 717 | 31 | 4.32 | 709 | 74 | 10.44 |
| **80 +** | 50529 | 13996 | 27.70 | 48320 | 10172 | 21.05 | 1430 | 208 | 14.55 | 227 | 50 | 22.03 | 485 | 117 | 24.33 |
| **TOTAL** | **199470** | **25215** | **12.64** | **210406** | **16517** | **7.85** | **44672** | **1023** | **2.29** | **6737** | **93** | **1.38** | **10765** | **247** | **2.31** |

Figure 1 compares the patterns of age-specific case proportions across the five countries. As can be seen, the patterns for Spain and Italy are skewed disproportionately towards older ages, while those for China, Australia and South Korea follow a relatively normal distribution. Table 2 shows that using the case distributions by age from South Korea as the standard, the age-adjusted CFR for Italy comes down to less than twice that of the South Korea, as compared to the five-fold difference in crude values. Also, the case-age adjusted CFR for Spain is now not much higher than the South Korean reference CFR, whereas the crude CFR was about three times the reference value. However, this difference remains statistically significant (see footnote to Table 2). Hence, the higher crude CFRs in Italy and Spain are largely influenced by age-structure, but the adjusted CFRs in both these populations indicate that there are other factors that could account for their higher values in relation to the CFR for South Korea, including the health service availability and readiness to respond to the surge in COVID 19 patient load from the epidemic. In contrast, the Australian adjusted CFR is the lowest of all countries when based on the South Korean standard, and this suggests a better case management program for diagnosed COVID 19 cases in Australia.

**Table 2:** Age standardised COVID 19 case fatality rates for study populations from different standards

| Country | Case population distribution standards |  |  |  |  |
| --- | --- | --- | --- | --- | --- |
|  | Italy | Spain | China | Australia | South Korea |
| Italy | **12.64** | 11.86 | 5.69 | 5.51 | 4.42 |
| Spain | 8.42 | **7.85** | 3.27 | 3.34 | 2.87 |
| China | 5.69 | 5.33 | **2.29** | 2.28 | 2.01 |
| Australia | 6.39 | 5.84 | 1.3 | **1.38** | 1.42 |
| South Korea | 8.29 | 7.66 | 2.45 | 2.55 | **2.31** |

Bold numbers indicate crude case fatality rates
Shaded areas indicate CFRs for Italy are $\approx 1.5$ to 2 times that of South Korea from all standards
Confidence intervals (95%) for the last column are Italy, 4.34 - 4.51; Spain, 2.79 - 2.94; China, 1.90 - 2.12; Australia, 1.16 – 1.67; South Korea, 2.08 - 2.54.

**Figure 1.**
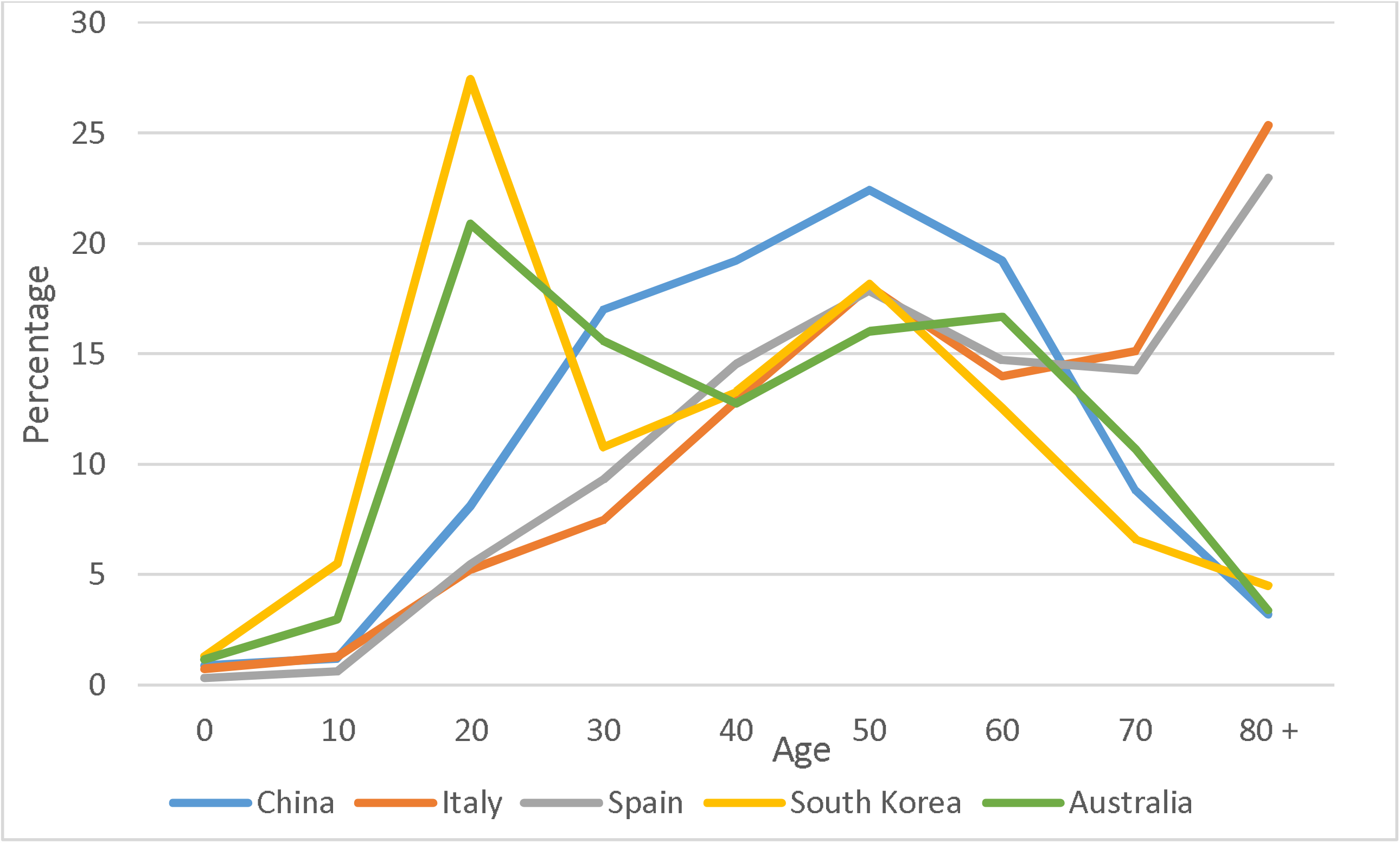
Age distribution of COVID 19 cases from study populations

## Discussion

Marked variations in crude CFRs have been a matter of concern for understanding the patterns of COVID 19 epidemic in different populations. Our results show that adjustment for age largely equalizes such variations, with the exception of the two-fold residual difference between the adjusted CFR for Italy (4.41%) and the South Korean reference value (2.31%). Standardization is a known technique to adjust for variations in age distributions of exposure across different populations, in order to derive comparable outcome measures. The observed crude CFR is not essentially a population based measure, since it is based on a case age distribution that does not necessarily mirror population age distributions. Hence, we adopted a novel approach of using age-distribution of confirmed cases in a population as the reference structure for standardization of the CFR. A reference case age distribution based on a sound screening, testing, notification and contact tracing program could subsume issues such as the influence of diagnostic test availability and strategies for case detection on the eventual numbers and distribution of COVID cases in different populations. Therefore, we recommend the South Korean distribution as the reference standard, since it is based on a rigorous notification and contact tracing intervention, and without any transmission suppression program. (16, 17)

Since the risk of severe disease or death after COVID-19 is now well known to increase with age, our results suggest that differences in the CFR should be taken to infer COVID 19 mortality differentials across populations only after adjusting for age structure of cases. Our inference regarding the influence of case age structure on CFR matches findings from another recent study, which used a decomposition method to evaluate this phenomenon.(18) The decomposition analysis derives a measure of the proportion of difference between the crude CFRs of study and reference populations that could be attributed to variations in the case age-structure of the study population, and the proportion due to other factors. However, these decomposed differences could vary across study populations due to their varying case age structures, even if their crude CFRs are similar. This technique is similar in concept to indirect standardization, and the derived measures can be used to infer age structure related mortality differentials between individual study and reference populations. However, these measures cannot be used to compare the effects of case age structure on CFR across different study populations. In contrast, our method of direct standardization can be used to derive age-adjusted measures of CFR that are comparable across all study populations.(19) Hence, given its ready interpretation, the directly standardized CFR could serve as an optimal measure for international comparisons of COVID 19 mortality.

In general, the CFR is an important measure during a developing epidemic, since there is inadequate information or exposure time to develop other population level epidemiological indicators. It is also of clinical importance as a measure, potentially at hospital level, to understand the burden from severe illness due to COVID on the health care system. The CFR could perhaps also serve as an indicator of care. For instance, even after adjusting for age structure, the high value of the standardised CFR seen in Italy possibly reflects the sudden increase in the burden on the health system, and the ensuing challenges in the quality of care delivered to severe cases. At the same time, there may be other patient related clinical drivers of higher CFRs, including nature of individual case co-morbidities, or frailty. In summary, the residual difference between the crude and directly standardised CFRs could represent the true health and health system related component of mortality differentials between populations.

As the pandemic matures, all countries will develop modalities for case detection, testing for disease, notification and contact tracing. Hence, it is likely that more reliable measures of incidence could be available, at least through a case definition for symptomatic individuals. Similarly, there would be concomitant improvements in accuracy of ascertainment of COVID mortality, to use general population based mortality indicators to monitor outcomes. In many developing countries, there is also a need to strengthen national mortality statistics programmes to support better epidemic surveillance.(20) Till then, CFRs might remain the main epidemiological measure; and case age-standardization could help improve international comparability of the progress of the pandemic, and guide disease prevention, control, and disease management strategies.

## Data Availability

All data used for this manuscript is available in the public domain, and appropriate references along with weblinks to data sources have been provided in the bibliography

## Notes

### Competing Interest Statement

The authors have declared no competing interest.

### Funding Statement

No external funding was received for preparing this manuscript

